# Consistency of left ventricular ejection fraction measurements in the early time course of STEMI

**DOI:** 10.1101/2023.01.13.23284539

**Authors:** Lilyana Georgieva, Fabian T. Nienhaus, Sebastian Haberkorn, Ralf Erkens, Amin Polzin, Patricia Wischmann, Rojda Ipek, Kian Marjani, Aikaterini Christidi, Michael Roden, Christian Jung, Florian Bönner, Malte Kelm, Stefan Perings, Mareike Gastl

## Abstract

**Background:** Assessment of left ventricular (LV) function and volume after ST-segment elevation myocardial infarction (STEMI) is recommended to guide clinical decision within and after hospitalization. Early after STEMI, initial LV reshaping and hypokinesia may affect analysis of LV function. A comparative evaluation of left ventricular ejection fraction (LVEF) and stroke volume (SV) by different imaging modalities to assess LV function early after STEMI has not been performed so far.

**Methods:** LV function was assessed by LVEF and SV using serial imaging within 24h and 5 days after STEMI with cineventriculography (CVG), 2-dimensional echocardiography (2DE), 2D and 3D cardiovascular magnetic resonance (2D/3D) in 82 patients. Respective parameters were compared between modalities and to 3D gold standard CMR.

**Results:** 2D analyses of LVEF using CVG and 2DE as well as 2D CMR yielded uniform results within 24h and 5 days of STEMI. SV assessment between CVG and 2DE at day 1 after STEMI was comparable, whereas values for SV were higher using 2D CMR on all occasions (p<0.01 all). This was due to higher LVEDV measurements. LVEF by 2D versus 3D CMR was comparable, 3D CMR yielded consistently higher volumetric values. This was not influenced by infarct location or infarct size.

**Conclusions:** Early after STEMI, 2D analysis of LVEF yielded robust results across all imaging techniques implying that CVG, 2DE, and 2D CMR can be used interchangeably in this setting. SV measurements to assess cardiac function differed substantially between imaging techniques due to higher intermodality-differences of absolute volumetric measurements.

## Introduction

Due to left ventricular (LV) reshaping and segmental hypokinesia, a serial determination of left ventricular function by ejection fraction (LVEF) and stroke volume (SV) is clinically relevant in ST-segment elevation myocardial infarction (STEMI). This is to monitor early time course of initial remodeling, risk assessment, and efficacy of therapeutic interventions [1,2]. Guidelines recommend that patients after myocardial infarction receive LV function assessment before hospital discharge [3]. In the early phase of acute myocardial infarction (AMI) immediate LV reshaping and segmental hypokinesia challenge exact imaging and analysis of LV function. Therefore, the analysis of LVEF and SV may yield distinct results, because they cover different aspects of LV function in relation to ventricular reshaping [4]. Available imaging techniques include two-dimensional unenhanced echocardiography (2DE), which is the most widely used technique due to its ease of use and availability. Without delaying reperfusion, cineventriculography (CVG) may help to guide decision making during primary percutaneous coronary intervention (pPCI). Cardiovascular magnetic resonance (CMR), the gold-standard for assessing cardiac volumes, provides additional prognostic information including indices of LV remodeling, infarct size (IS) and area of edema [2,5,6].

Length of hospital stays for STEMI patients have become shorter over the last decades [7]. STEMI patients are discharged from regular wards after 48h. During this acute phase of AMI, distinct imaging modalities are frequently applied to monitor LV function early pending on availability in the catheter laboratory, the intensive and coronary care unit, and regular wards. A follow-up is usually performed by an outpatient cardiologist using 2D imaging as the mode of choice.

There is a gap in knowledge whether or not the available imaging modalities in the early phase after STEMI provide uniform LVEF and SV assessment and whether 2D analyses are comparable to 3D analyses. This raises the important question whether or not analysis of parameters for measurement of LV function can be applied interchangeably across all imaging modalities. In addition, there are no data available on serial comparison of 2D vs 3D mode of analysis in the early phase of AMI. The hypothesis of this study was that serial assessment of LVEF in contrast to SV yields robust results across applied imaging modalities independent in the early phase of STEMI.

## Methods

### Study design

The aim of this study was to evaluate interchangeability of CVG, 2DE and 2D CMR for the assessment of LVEF and SV in the early time course of STEMI and whether LV reshaping may affect these measurements. Analyses were validated against the gold standard of LV function (3D CMR). Data were obtained from the systemic organ communication in STEMI cohort (SYSTEMI, Identifier NCT03539133). SYSTEMI is an open-end prospective cohort study for the prospective and systematic collection of clinical research data from STEMI patients. Ethics approval was given by the local ethics committees (Heinrich-Heine-University, Düsseldorf, Germany) complying with the Declaration of Helsinki. SYSTEMI aims to identify targetsthat are critical for the acute and subacute response after acute myocardial ischemia and most likely determine LV function and remodelling with consecutive heart failure endpoints [8]. Data from a serial assessment of global LV function at different time points in the context of SYSTEMI is important to investigate pharmacological interventions on LV function in the early time course of STEMI.

### Imaging sub-study design

Inclusion criterion of the present imaging sub-study as part of SYSTEMI was serial imaging with CVG, 2DE and CMR within 24h and 5 days after STEMI (*Fig 1A*). LV functional parameters of interest were LVEF and SV. Those parameters were calculated based on LVEDV and LVESV measurements. Analyses of CVG, 2DE and 2D and 3D CMR were performed by different readers in a blinded fashion during clinical routine or for research purposes. As only one reader was trained for CVG analyses, additional interobserver agreement was performed for CVG.

**Figure 1.**
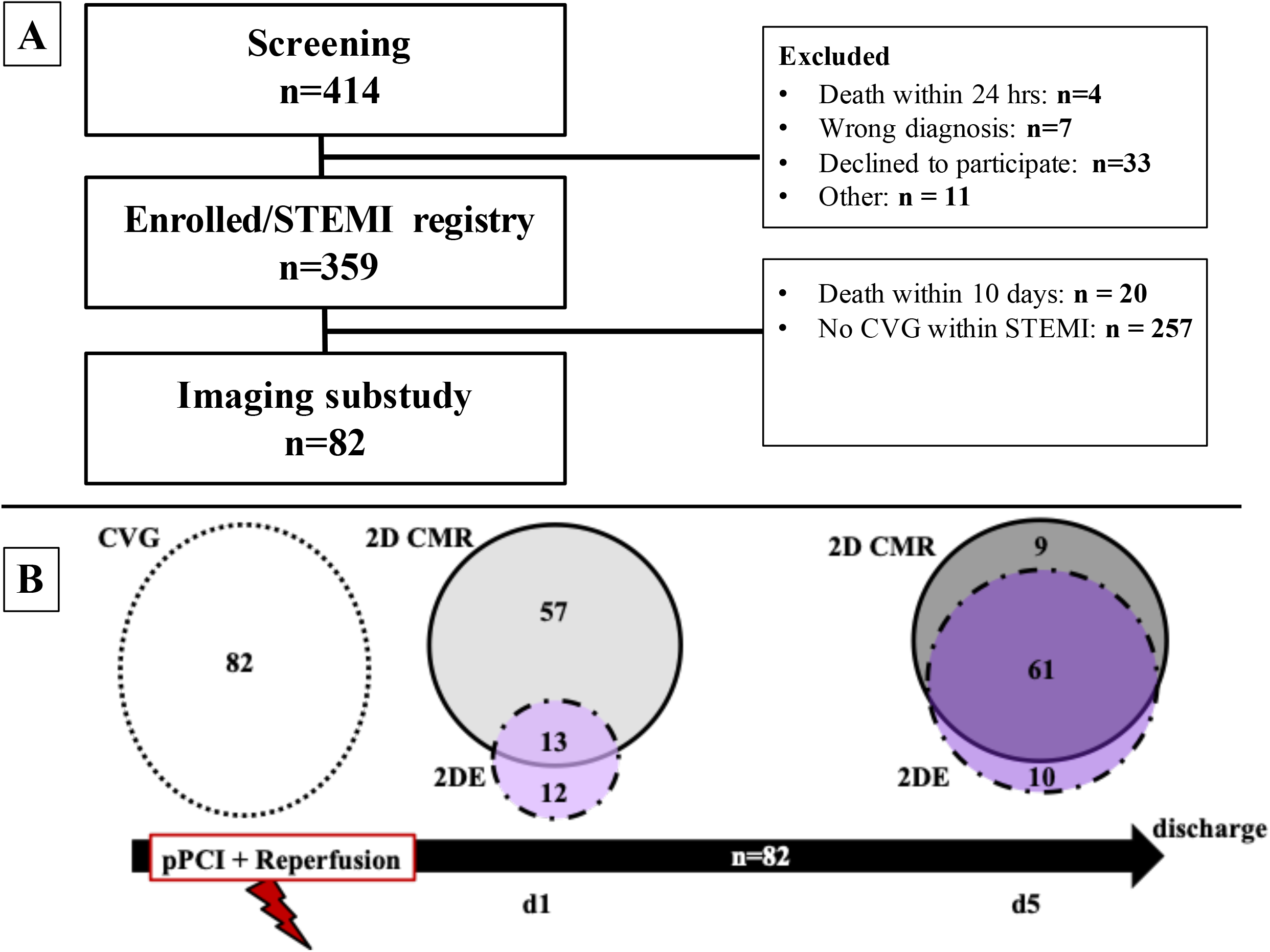
Consort diagram and timeline. A final 82 patients with STEMI and serial imaging in the early time course of STEMI were included (A). CVG (dotted circle) was performed within pPCI, 2DE (striped circle) and CMR (continuous circle) were performed 1 and 5 days after pPCI (B). Overlapping modalities per patient are indicated in white numbers. 2DE: 2D echocardiography; CVG: cineventriculography; CMR: cardiovascular magnetic resonance; EF: ejection fraction; LV: left ventricular; pPCI: primary percutaneous coronary intervention; STEMI: ST-segment elevation myocardial infarction; SV: stroke volume.

### Cineventriculography

Standard biplane CVG was obtained using a 30° right anterior oblique (RAO) projection and a 60° left anterior oblique (LAO) projection with the injection of at least 20 mL of contrast medium (Accupaque 350, GE Healthcare Büchler GmbH & Co. KG) at a flow rate of 14 mL/s using 6F pigtail catheters. In 30 patients, monoplane CVG in RAO projection was available. CVG was performed within catheterization for pPCI.

LV volumes and function were determined using biplane Simpson’s method for all patients according to well-defined standards using the LV analysis module included in the CAAS II software (Pie Medical, Maastricht, The Netherlands). Semi-automatic boarder tracking was used in end-diastole and end-systole in the frames with the largest and smallest ventricular silhouettes with succeeding calculation of LV parameters. Contours were amended, if necessary.

### Transthoracic echocardiography

2DE was obtained using a commercially available ultrasound scanner (Vivid E95, GE Vingmed, Horten, Norway) in the early hours (median 46.6 hours, interquartile range (IGR) 27.4 – 53.9 hours) and 5 days after pPCI (median 108.6 hours, IQR 89.6 – 128.8 hours) (*Fig 1B*). For endocardial border definition, images were optimized for each patient by modulation of transmit power, gain, focus, and dynamic range. Apical four-chamber, two-chamber views as well as a full 3D volume data set were acquired. The patients were investigated in the left lateral recumbent position, if possible. All data sets were analysed on-side using dedicated software as established on the ultrasound scanner, according to current standards and after formal training [9]. LV function and volumes were determined by manually tracing end-systolic (smallest LV shape) and end-diastolic endocardial borders (largest LV shape) in respective apical four-chamber and two-chamber views. Simpson’s method for biplane assessment was used.

### Cardiovascular magnetic resonance

CMR imaging was performed on a 1.5T MR imaging system (Achieva, Philips Healthcare, Best, The Netherlands) using a 32-channel phased array coil in the early hours (median 23.1 hours, IQR 16.2 – 33.1 hours) and 5 days (median 120.2 hours, IQR 104.3 – 130.6 hours) after pPCI. Standard cine steady-state-free precession (SSFP) images in standard long-axis geometries (two-, and four-chamber view) on all occasions was performed. Due to time saving reasons, acquisition of short-axis SSFP-images with full ventricle coverage from basis to apex was only performed on day 5 after pPCI. Sequence parameters of all scans can be found in the *S1 File*.

Certified CMR evaluation software was used for all analyses (CVI42, Circle Cardiovascular Imaging Inc., Calgary, Alberta, Canada or Sectra Workstation IDS7, Version 19.3.6.3510). Automatic calculation with manual adjustment, if necessary, of all LV volumes and functions was performed from short-axis SSFP images to obtain 3D datasets. As short-axis orientation with full ventricle coverage was not available in the first day after pPCI and for a better comparison to CVG and 2DE, additional assessment of LV parameters in all CMR measurements was performed using a biplane module. Automatic endocardial detection was performed in 2D four-chamber and two-chamber views after manual definition of the LV extent in end-diastolic and –systolic frames of both views. Contours were amended manually, if necessary, and papillary muscles or trabeculations were allocated to the LV cavity.

### Statistical analysis

Statistical analysis was performed using SPSS 25.0 (SPSS Inc., Chicago, IL, US). Unless otherwise stated, continuous variables are presented as mean□±□standard deviation (SD). The Shapiro-Wilk test was used to test normal distribution. Categorical variables are reported as percentage. Data between the different imaging modalities were analysed by the paired-samples t-test for normally distributed data and the Wilcoxon signed-rank test for not normally distributed data. The Fisher’s exact test was used to examine significant differences between nominal classifications. Pearsons or Spearman correlation were used to correlate the different parameters between the modalities and dimensions for normally or not-normally distributed data respectively. Coefficients of variation (CoV) were calculated during Bland–Altman analyses as the SD of the differences divided by the mean values. Intraclass correlation coefficients (ICC) were assessed using a model of absolute agreement for interobserver agreement. There was excellent agreement when the ICC was > 0.74, good when ICC = 0.60–0.74, fair when ICC = 0.40–0.59, and poor when the ICC was < 0.41 [10].

To determine an impact of infarct territory on a subset of patients (2D versus 3D CMR measurements), patients were divided into anterior and non-anterior STEMI. Respective statistical tests were performed between the different modalities and dimensions again. Two-tailed p-values below 0.05 are considered statistically significant.

## Results

### Baseline characteristics

As shown in *Figure 1A*, 359 patients were enrolled for SYSTEMI from 2018 to 2020. Complete phenotyping according to the SYSTEMI protocol was provides in 359 patients. Serial imaging with CVG as well as 2DE or 2D CMR within 24h after STEMI and 5 days after pPCI was conducted in 82 patients. Due to the all-comer design, not every patient had both, 2DE and 2D CMR, within the first hours after STEMI. Detailed numbers in respective analyses are visualized in black in *Figure 1B*. Patients with overlapping image modalities are given in white numbers. Baseline characteristics of the population as compared to the whole study cohort of SYSTEMI are presented in *Table 1*. There was a trend that patients included in the serial imaging substudy of SYSTEMI were younger and had shorter door-to-balloon time associated with lower Killip class, better renal function, and reduced inflammatory parameters, as compared to the entire cohort.

**Table 1.** Patient characteristics. p<0.05 indicates a significant difference between parameters of the imaging substudy compared to the whole cohort of SYSTEMI using unpaired two-tailed t-tests or Mann-Whitney U-test (marked with #) for not normally distributed data. Fisher’s exact or χ2 test was used to examine significant differences between nominal classifications.

|  | Imaging substudy<br>N = 82 | STEMI registry<br>N = 277 | p-value |
| --- | --- | --- | --- |
| Age, years | 60 [54 70] | 63 [54 74] | 0.120 |
| Male, n[%] | 63 [77] | 199[72] | 0.399 |
| BMI, kg/m<sup>2</sup> | 25.4 [24 28.3] | 26.3 [24.0 29.4] | 0.061 |
| **<u>Cardiovascular risk factors</u>** |  |  |  |
| Current smoking, n[%] | 43[52] | 127[47] | 0.652 |
| Hypertension, n[%] | 57[70] | 191[70] | 1.000 |
| Hypercholesterolemia, n[%] | 44[54] | 122[45] | 0.166 |
| Diabetes mellitus, n[%] | 23[28] | 74[27] | 0.888 |
| Previous infarction, n[%] | 13[16] | 41[15] | 0.861 |
| Previous PCI, n[%] | 14[17] | 43[16] | 0.731 |
| Previous CABG, n[%] | 1[1] | 6[2] | 1.000 |
| SBP, mmHg | 149±27 | 144±31 | 0.184 |
| DBP, mmHg | 91 [82 100] | 87 [77 100] | 0.193 |
| Heart rate, bpm | 78 [66 93] | 80 [66 97] | 0.419 |
| Chronic kidney disease |  |  |  |
| Creatinine clearance, ml/min | 86 [75 99] | 79 [60 95] | **0.003** |
| Serum creatinine, µmol/l | 0.86 [0.77 1.02] | 0.97 [0.8 1.2] | **0.007** |
| **<u>Procedural data</u>** |  |  |  |
| Killip class on admission |  |  | **<0.001** |
| 1, n[%] | 64[80] | 122[47] |  |
| 2, n[%] | 11[14] | 31[12] |  |
| 3, n[%] | 2[3] | 13[5] |  |
| 4, n[%] | 3[4] | 92[36] |  |
| No. of diseased vessels |  |  | 0.276 |
| 1, n[%] | 15[18] | 40[15] |  |
| 2, n[%] | 15[18] | 36[13] |  |
| 3, n[%] | 52[63] | 198[73] |  |
| Infarct related artery |  |  | 0.297 |
| Left anterior descending | 40[49] | 124[45] |  |
| Right coronary artery | 29[35] | 119[44] |  |
| Left circumflex | 13[16] | 30[11] |  |
| Max. stent length, mm | 16 [15 20] | 16 [15 18] | 0.920 |
| Thrombectomy | 6[7] | 35[13] | 0.236 |
| Drug-eluting stent | 77[94] | 260[95] | 0.788 |
| TIMI flow grade post-PCI |  |  | 0.977 |
| II | 5[6] | 20[8] |  |
| III | 73[90] | 230[88] |  |
| **<u>Laboratory parameters</u>** |  |  |  |
| CK max. [U/L] | 970 [520 2275] | 1266 [520 2873] | 0.065 |
| CRP max. [mg/dL] | 2.8 [1.2 6.3] | 7.1 [2.3 17] | **<0.001** |
| NT-proBNP max [pg/mL] | 1256 [719 2446] | 2029 [726 4786] | **0.030** |
| Troponin T [ng/L] | 2908 [981 6755] | 3136 [1114 7508] | 0.339 |

ACE, angiotensin-converting enzyme; BMI, body-mass-index; CABG, coronary artery bypass graft; CK, creatine kinase; CRP, C-reactive protein; DBP, diastolic blood pressure; NT-proBNP, brain
natriuretic peptide; PCI, percutaneous coronary intervention; SBP, systolic blood pressure; STEMI, ST-segment elevation myocardial infarction; TIMI, thrombolysis in myocardial infarction.

### Analyses of LV function by LVEF

All imaging modalities were available for analyses in a 2D view on both occasions after STEMI. The rationale was to test for robust LVEF measurements using CVG, 2DE and 2D CMR independent of the time point of measurement. Results indicated that LVEF assessment can be performed independent of the imaging modality and timing after STEMI. LVEF values were similar when determined by either CVG, 2DE, or 2D CMR in the early time course of STEMI (CVG vs. 2D CMR: 54±9 vs. 54±10%, 2DE vs. 2D CMR: 52±8 vs. 52±10%) (*Fig 2A*). This was mirrored comparing 2DE and 2D CMR 5 days after STEMI (2DE vs. 2D CMR: 56±10 vs. 55±10%) (*Fig 3A*). LVEF values by CVG, 2DE and 2 D CMR were significantly correlated with a good CoV (CoV LVEF CVG vs. 2DE: 12%; CVG vs. CMR 10%, 2DE vs. 2D CMR 11%) (*Suppl. Fig 1, Suppl. Fig 2A, Suppl. Table 1*).

**Figure 2.**
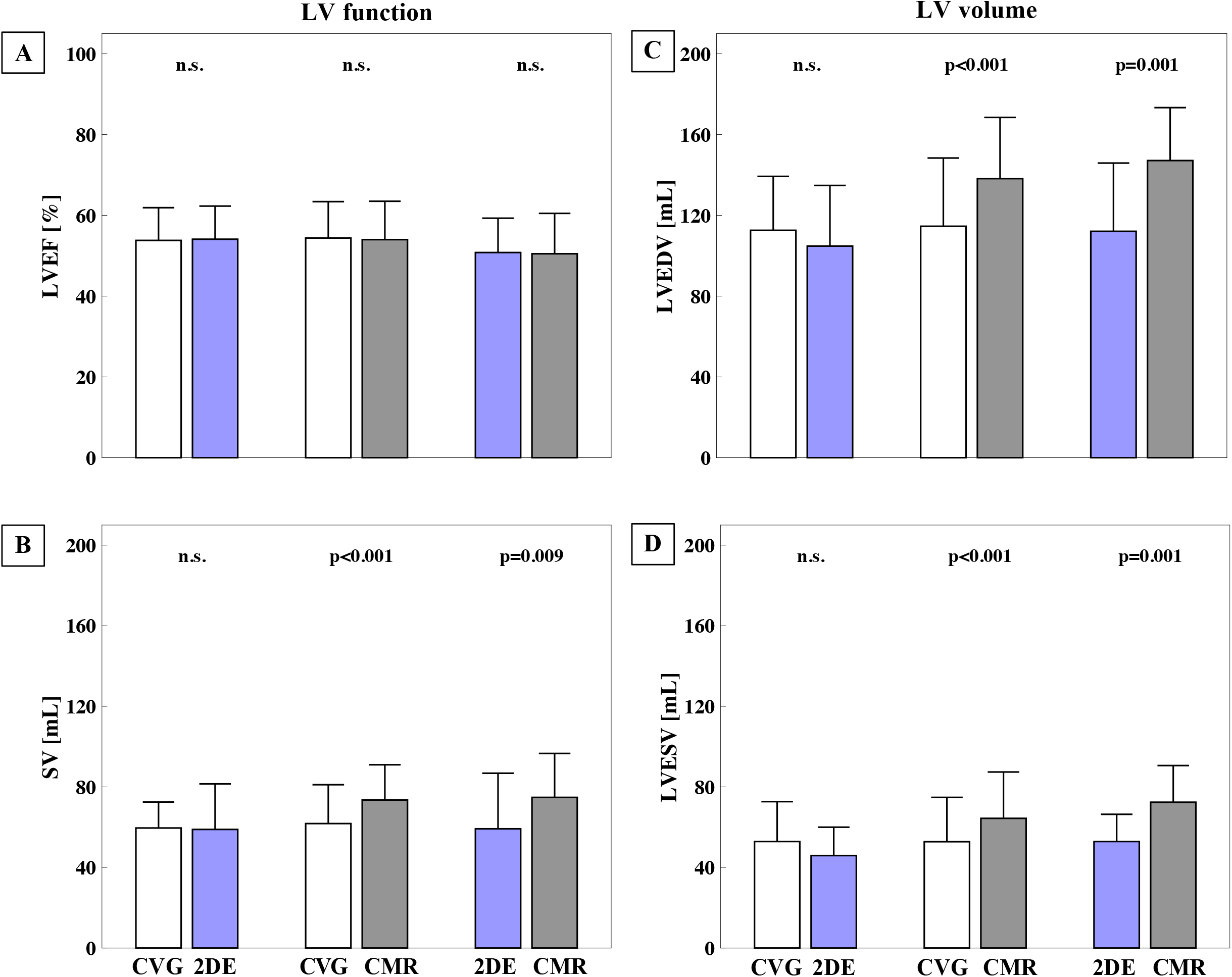
Consistency of 2D LVEF assessment within 24h of STEMI across imaging modalities. Descriptive statistics (mean±SD) are given for LV function (A, B) and volume (C, D). LVEF assessment was robust between all applied imaging modalities (. SV values were not interchangeable due to lower volumes by either CVG or 2DE as compared to 2D CMR (A). SV was comparable between CVG and 2DE (B). p<0.05 indicates a significant difference using paired-samples t-test for normally distributed data. 2DE: 2D echocardiography, CVG: cineventriculography; CMR: cardiovascular magnetic resonance; EDV/ESV: end-diastolic/-systolic volume; EF: ejection fraction; LV: left ventricular; SD: standard deviation; STEMI: ST-segment elevation myocardial infarction; SV: stroke volume.

**Figure 3.**
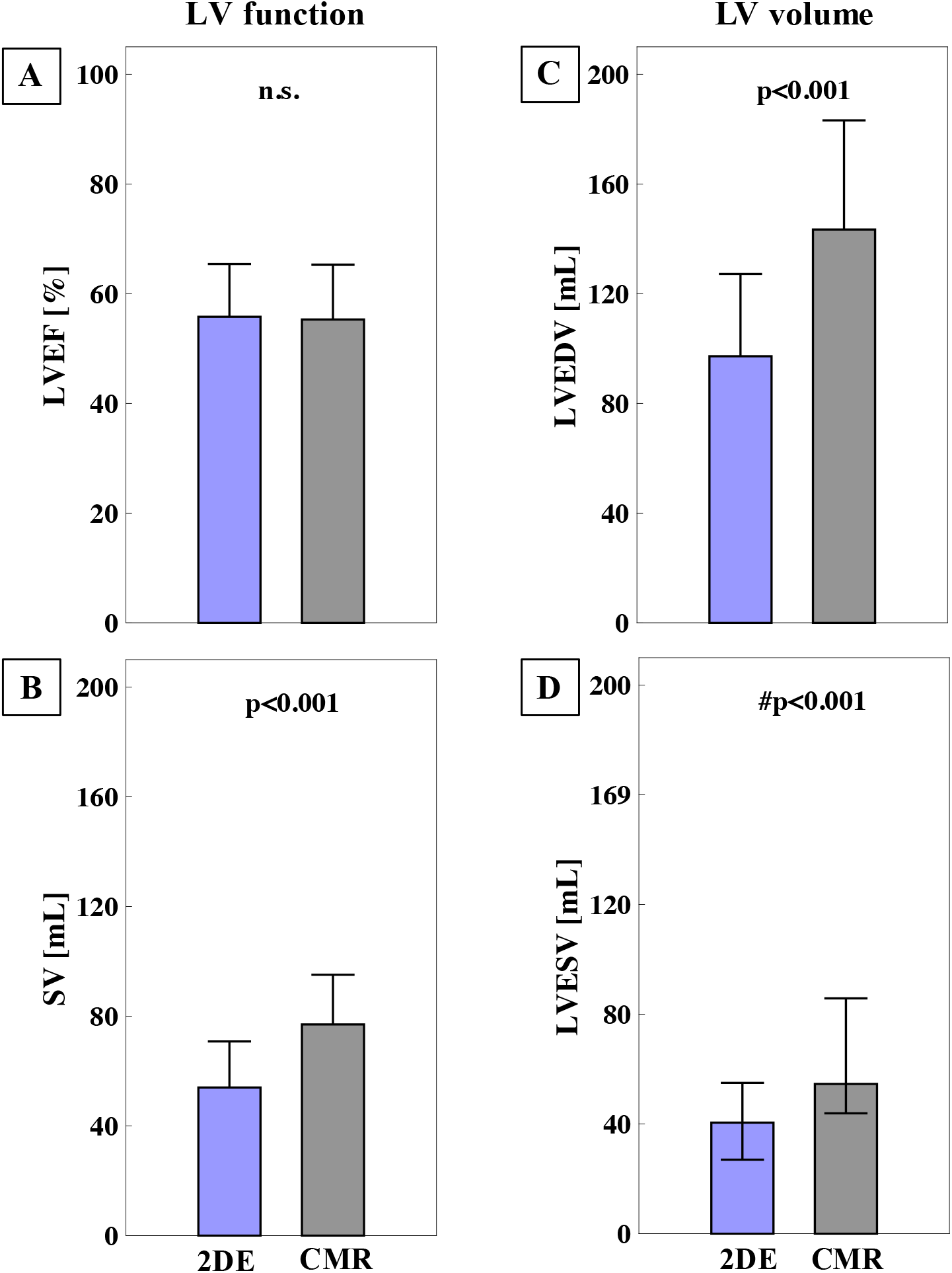
Consistency of 2D LVEF assessment at 5 days after pPCI across imaging modalities. Descriptive statistics (mean±SD) are given for LV function and volume. LVEF assessment was reliable between 2DE and 2D CMR (A). SV values (B) were not interchangeable due to lower volumes by 2DE as compared to 2D CMR (C, D). p<0.05 indicates a significant difference using either paired-samples t-test for normally distributed data. 2DE: 2D echocardiography, CMR: cardiovascular magnetic resonance; EDV/ESV: end-diastolic/-systolic volume; EF: ejection fraction; LV: left ventricular; SD: standard deviation; STEMI: ST-segment elevation myocardial infarction; SV: stroke volume; pPCI: primary percutaneous coronary intervention

### Analyses of LV function by SV

SV is an important descriptor of cardiac function independent from LVEF and was examined between imaging modalities and all time-points after STEMI separately from LVEF measurements. Results indicated that different imaging modalities cannot be used interchangeably for SV assessment. SV values as analysed from 2D CMR were higher as compared to values obtained from CVG and 2DE on both occasions after STEMI (CVG vs. 2D CMR: 61.7±19.3 vs. 73.7±17.5mL, p<0.001, 2DE vs. 2D CMR: 52.9±14.1 vs. 77.6v±19.9mL, p=0.009). This was attributable to higher differences in LVEDV (difference CVG vs. 2D CMR 23.6±32.2mL, difference 2DE vs. 2D CMR 36.4±33.2mL) as compared to LVESV (difference CVG vs. 2D CMR 11.7±17.2mL, p<0.001, difference 2DE vs. 2D CMR 19.9±18.6mL, p=0.01) (*Fig 2B and 3B*). SV values by CVG compared to 2DE were similar. Correlations results underlined differences of SV measurements between all modalities (*Suppl. Fig 2B, Suppl. Table 1*).

As LV function was determined on the basis of LV volumes, LVEDV and LVESV were analysed as well for complimentary reasons. Measurements for LVEDV and LVESV showed higher values in 2D CMR compared to CVG and 2DE at both times after STEMI (LVEDV CVG vs. 2D CMR: 114.6±33.8 vs. 1338.2±30.8mL, p<0.001, LVEDV 2DE vs. 2D CMR: 114.4±34.4 vs. 150.4±24.1mL, p=0.001, LVESV CVG vs. 2D CMR 52.8±22.0 vs. 64.4±23.0 mL, p<0.001, LVESV 2DE vs. 2D CMR: 52.9±14.1 vs. 72.8±18.8mL, p=0.001) (*Fig 2C/D and Fig 3C/D*). Values between CVG and 2DE were comparable. This was underlined by correlation analyses (*Suppl. Fig 2C/D, Suppl. Table 1*). There was excellent reproducibility of LV function by LVEF and SV as well as LVEDV and LVESV using CVG as imaging method (ICC from 0.86 for LVEF to 0.96 for LVEDV).

### 2D versus 3D CMR analyses of LVEF and SV

In a subgroup of patients (n=69), 3D CMR was chosen as reference gold standard for comparing LV function assessment. 3D analyses demand for more acquisition time. Therefore, 3D CMR measurements were only performed 5 days after pPCI due to the instability of critically ill patients within the first 24h after STEMI. LVEF values were not different between 2D and 3D CMR dimensions (2D vs. 3D CMR: 56±10 vs. 55±10%) (*Fig 4, Suppl. Table 2)*. SV analyses were higher in the 3D mode (2D vs. 3D CMR: 77.4±17.8 vs. 86.8±18.3mL) (*Fig 4, Suppl. Table 2*). This was again attributable to higher differences in LVEDV (LVEDV 2D vs. 3D CMR: 143.6±38.3 vs. 163.4±38.0mL, p<0.001, difference 19.8±17.3 mL) than LVESV values (LVESV 2D vs. 3D CMR: 66.2±30.3 vs. 76.6±33.1mL, p<0.001, difference 10.5±11.9 mL, p<0.001). Therefore, SV values cannot be used interchangeably when analysed by 2D CMR or 3D CMR.

**Figure 4.**
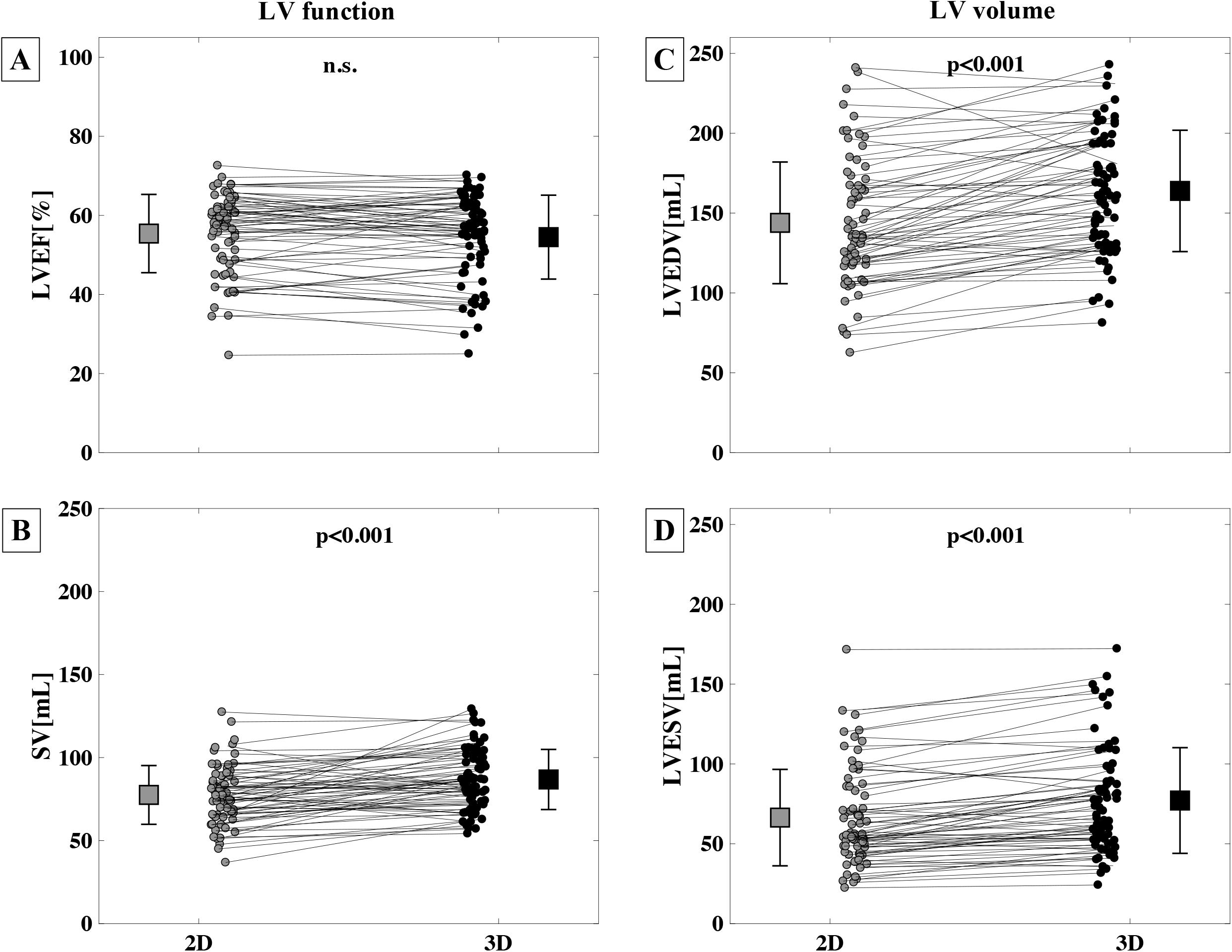
Consistency of LVEF assessment at 5 days after pPCI across 2D and 3D CMR dimensions. Descriptive statistics (mean±SD or median [IQR, marked with #]) are given for comparisons of LVEF (A), SV (B), LVEDV (C), LVESV (D) between CMR dimensions. LVEF assessment remained reliable between CMR dimensions, a change in CMR dimensions between SV assessments should be avoided. p<0.05 indicates a significant difference using either paired-samples t-test for normally or Wilcoxon signed-rank test (marked with #) for not normally distributed data. CMR: cardiovascular magnetic resonance; EDV/ESV: end-diastolic/-systolic volume; EF: ejection fraction; LV: left ventricular, PCI: primary percutaneous coronary intervention; SD: standard deviation; SV: stroke volume

### Influence of infarct territory and infarct size

To reveal whether the infarct territory and infarct size influence 2D versus 3D measurements due to geometric assumptions of 2D measurements, patients were divided into anterior versus lateral/inferior infarct location. Only examining patients with anterior STEMI (n=36), results were not different as compared to the whole cohort. LVEF was still comparable between 2D and 3D CMR (p=0.269) whereas SV values were higher in the 3D mode (2D vs. 3D: 75.59±18.69 vs. 83.04±17.84 mL, p=0.001). In non-anterior STEMI (n=24), results were the same (LVEF: p=0.121; 2D vs. 3D SV: 77.38±18.09 vs. 88.61±18.84 mL, p<0.001).

Differences between 2D and 3D CMR were correlated to the IS 5 days after STEMI. IS did neither correlate to the differences in LVEF nor in SV. There was no correlation for LVEDV and LVESV differences in dimensions.

## Discussion

Using a serial, cross-modality imaging approach with CVG, 2DE and 2D CMR, this study showed robust LVEF assessment independent of the imaging modality in the early time course after STEMI. The uniform LVEF measurement was also detectable 5 days after STEMI. When using CMR, assessment of LVEF values were independent from the imaging dimension, either 2D or gold standard 3D. CMR showed higher absolute values when measuring SV compared to CVG and 2DE. This was due to higher absolute differences in LVEDV measurements between imaging modalities. When interpreting LV function after STEMI, LVEF measurements consider changes in cardiac output and remodelling through its calculation via normalization of the SV to the LVEDV. In contrast to SV measurements, differences between imaging modalities may therefore be compensated when using LVEF.

### Impact factors on the assessment of LV function early after STEMI

Imaging of LV function and volume early after AMI allows to evaluate risk assessment and the impact of therapeutic interventions, designed to preserve cardiac function and predict long-term remodelling. However, the assessment of LV function after STEMI is being complicated by hypokinesia in non-affected segments and myocardial remodelling [4].LV function can be assessed by LVEF and SV. As LVEF is calculated by dividing SV through LVEDV, changes in LVEF may be due to changes in either SV, LVESV, LVEDV or a combination of those [4,11]. Therefore, LVEF is affected by preload, afterload and contractility and reflects both, changes in LV function and LV remodeling [12]. This is important in the setting of AMI with LV reshaping. Both, LVEDV and LVESV may be increased. This preserves SV, but decreases LVEF. In other circumstances, SV becomes an important parameter for LV function assessment. In heart failure with preserved ejection fraction, despite high-normal LVEF a low SV is inadequate to generate cardiac output leading to heart failure symptoms even when LVEF is considered normal [13]. SV is therefore more likely to reflect cardiac output and not myocardial remodeling that is present after STEMI. The current results propose a uniform assessment of LVEF throughout different imaging modalities and at two time frames early after STEMI. The calculation of LVEF as a division of SV through LVEDV may balance differences in single parameters, but is more likely to reflect combined changes in cardiac output and remodeling early after STEMI [4].

### Cross-modality analyses of LV function early after STEMI

Due to a faster transition of STEMI patients from catheterization labs through coronary or intensive care units to normal wards and finally early discharge, different imaging modalities may be applied at short time slots. Outpatient cardiologists using echocardiography for LV function assessment may benefit from measurements comparable to in-hospital acquisitions. A 2D imaging approach is usually used for time saving reasons.

Existing guidelines recommend the assessment of LVEF after STEMI before hospital discharge [14]. Echocardiography is recommended as the preferred imaging technique. It is mostly used as 2DE mode. A possible advantage of CVG is its ease of acquisition within pPCI before reperfusion in the acute phase of AMI without delaying reperfusion and in addition to provide information on regional LV function in the guidance of coronary revascularization in multivessel desease. Calculation of LVEF by CVG requires the same geometric assumptions as made for 2DE. 2DE and CVG may both suffer from inadequate acoustic or X-Ray windows to display endocardial borders of LV function assessment. Foreshortening of the left ventricle by tangential cuts may further complicate defining the real LV apex. CMR can overcome some of these shortcomings, but it is more time consuming and not widely available. It is able to visualize endocardial borders with high resolution. Therefore, hypokinetic regions or regions with papillary muscles can better be distinguished. In addition, CMR it is able to provide information with additional prognostic relevance such as IS and edema extent [15]. There is a lack of knowledge about the comparison of LV function assessment using these imaging modalities in the acute setting of STEMI. In contrast to CCS no studies assessed comparatively the imaging analysis of LV function in the very early phase of STEMI.

The present study results showed comparable 2D LVEF assessment within the first day after STEMI. Uniform LVEF assessment could be reproduced 5 days after pPCI for STEMI in dependent of applied imaging modality implying that interchangeable use of these techniques in the early phase of AMI is feasible. In contrast, analysis of SV was not consistent over all imaging analyses. CVG and 2DE underestimated LVEDV and LVESV as compared to 2D CMR. This underestimation was most pronounced for LVEDV leading to differences in SV. Therefore, it is reasonable to assume that a 2D mode is a valid parameter for the detection of LVEF changes in the acute time course after STEMI independent of the imaging modality.

### 2D versus 3D analyses of LV function early after STEMI

The majority of imaging modalities that rely on biplane acquisition require inputs about the ventricle geometry for volume calculations [16]. Therefore, an irregularly shaped LV in the setting of STEMI may be missed due to assumptions of regularly shaped ventricles in mathematical models [4]. Contrast-administration or 3D acquisition techniques are suggested to improve comparability of LV function assessment [17,18]. In various clinical settings, 3D CMR is considered the gold standard for the assessment of LV function [4,19]. Therefore, in this study 2D was compared to 3D CMR in a subset of patients 5 days after STEMI for the first time. 3D CMR imaging is more time consuming than 2D CMR. In this study, 3D CMR did not improve accuracy of LVEF determination over 2D CMR. SV and LV volumes were higher in 3D measurements. As LVEF is calculated from SV and LVEDV and both values increased, this may balance the differences in single values. The differences in SV can be explained by an unequivocal increase of LVEDV compared to LVESV. The increase of SV and LV volumes using a 3D mode could not be attributed to an influence of the IS or the location of STEMI in which 2D CMR may miss distinct regions of the heart. Another possible explanation is the 3D coverage of the whole ventricle that may increase detection of papillary muscles or LV remodeling.

These results indicate that 2D CMR may reliably be used to assess LVEF after STEMI. LVEF is as well independent of the infarct location or LV remodeling.

## Limitations

From 359 enrolled STEMI patients in SYSTEMI, 82 patients were part of this sub-study. Transient instability of STEMI patients may affect the choice of the applied imaging technique,. As shock patients could not be included for imaging, especially CMR, LVEF values of this study’s STEMI patients are not impacted severely. Therefore, these results should not be applied for all critically ill patients after STEMI. Inter- and intraobserver variability have not been obtained in the present study for echocardiography and CMR. There is already extensive data on the variability of the different imaging modalities and dimensions showing a good ICC for these modalities [20,21].

## Conclusion

Within 1 and 5 days after STEMI, 2D analysis of LVEF, but not SV, yielded consistent assessment of LV function across all imaging modalities irrespective of the chosen time point. Consequently, CVG, 2DE and 2D CMR can be used interchangeably for LVEF assessment. Pending on the applied imaging analysis, values for SV may vary across imaging modalities in the early time course after STEMI. Both, LVEF and SV cover advantages and disadvantages for LV function description in the early time course after STEMI. As LVEF is calculated from SV and LVEDV it is more likely to simultaneously reflect cardiac output and initial LV remodelling.

## Data Availability

All data produced in the present work are contained in the manuscript

## Acknowledgments

We acknowledge the support of staff including Miss Stefanie Bensmann, Joy Dillenburg and Juliane Geisler at the Department for Cardiology, Angiology and Pulmonology, University Hospital in Düsseldorf, Germany. We also acknowledge the Clinical Trial Unit (CTU) and Moritz Gastl.

## Supplemental Figures

**Suppl. Figure 1. LVEF assessment reveals good correlations and COVs between CVG, 2DE and 2D CMR within 1 days after STEMI.**

Correlation measurements and Bland-Altman analyses were performed on LVEF measurements between the different imaging modalities. Analyses underlined robust LVEF measurements between CVG (A, B), 2DE (A, C) and 2D CMR (B, C).

r indicates the correlation values as obtained from Pearson correlation, r_s_ from Spearman correlation for not normally distributed data.

2DE: 2D echocardiography; CMR: cardiovascular magnetic resonance; CoV: coefficient of variance; EDV/ESV: end-diastolic/-systolic volume; EF: ejection fraction; LV: left ventricular; STEMI: ST-segment elevation myocardial infarction; SV: stroke volume.

**Suppl. Figure 2. LVEF assessment reveals good correlations and CoVs between 2DE and 2D CMR 5 days after STEMI.**

Correlation measurements and Bland-Altman analyses were performed on LV function (A, b) and volume (C, D) between 2DE and 2D CMR. Analyses underlined interchangeability of LVEF measurements between 2DE and 2D CMR 5 days after STEMI (A). Due to a high CoV and decreased correlation, SV comparisons were not as robust as LVEF measurements (B).

r” indicates the correlation values as obtained from Pearson correlation, r_s_ from Spearman correlation for not normally distributed data.

2DE: 2D echocardiography; CMR: cardiovascular magnetic resonance; CoV: coefficient of variance; EDV/ESV: end-diastolic/-systolic volume; EF: ejection fraction; LV: left ventricular; STEMI: ST-segment elevation myocardial infarction; SV: stroke volume. pPCI: primary percutaneous coronary intervention

## Supplemental Tables

**Suppl. Table 1.**
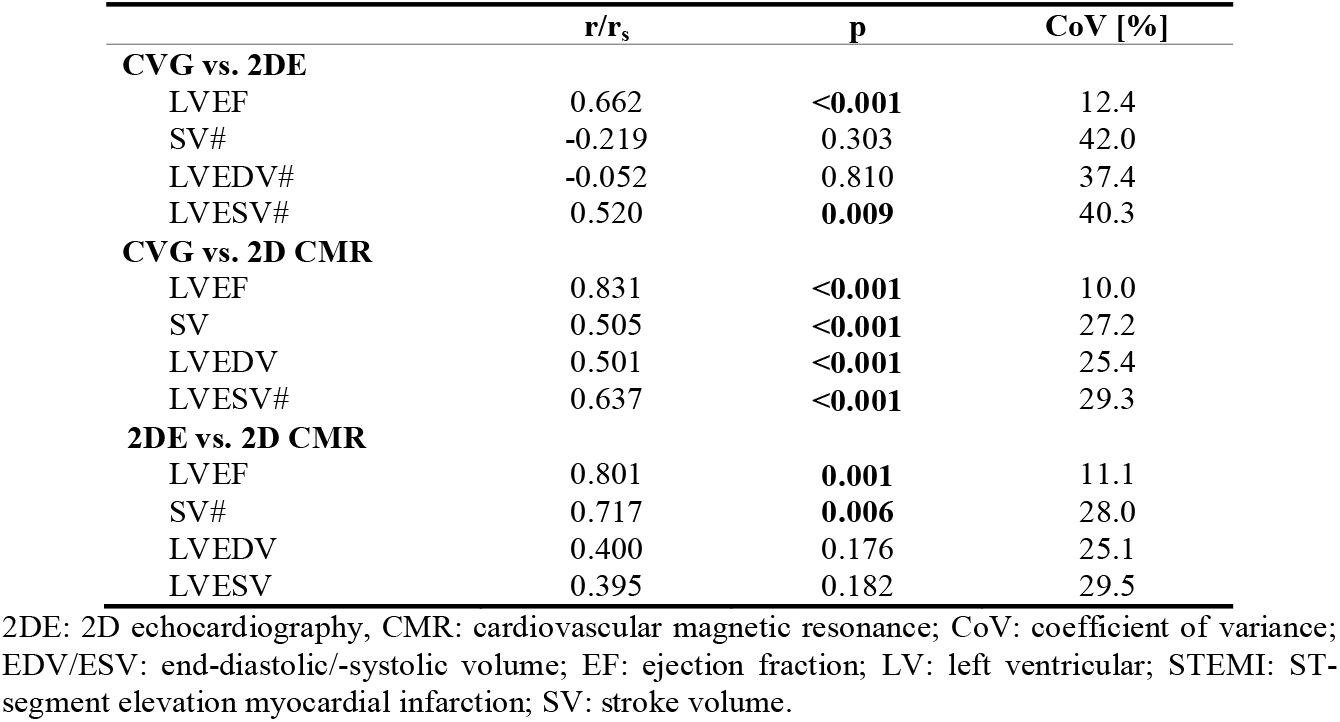
Correlation analyses and CoV between measurements of CVG, 2DE and 2D CMR within and at day 1 after reperfusion for STEMI. Results underlined that LVEF assessment can be performed using different imaging modalities. SV assessment should be performed within one modality. r indicates the correlation values as obtained from Pearson correlation or r_s_ from Spearman correlation (marked with #) for not normally distributed data.

|  | **r/$r_s$** | **p** | **CoV [%]** |
| --- | --- | --- | --- |
| **CVG vs. 2DE** |  |  |  |
| LVEF | 0.662 | **<0.001** | 12.4 |
| SV# | -0.219 | 0.303 | 42.0 |
| LVEDV# | -0.052 | 0.810 | 37.4 |
| LVESV# | 0.520 | **0.009** | 40.3 |
| **CVG vs. 2D CMR** |  |  |  |
| LVEF | 0.831 | **<0.001** | 10.0 |
| SV | 0.505 | **<0.001** | 27.2 |
| LVEDV | 0.501 | **<0.001** | 25.4 |
| LVESV# | 0.637 | **<0.001** | 29.3 |
| **2DE vs. 2D CMR** |  |  |  |
| LVEF | 0.801 | **0.001** | 11.1 |
| SV# | 0.717 | **0.006** | 28.0 |
| LVEDV | 0.400 | 0.176 | 25.1 |
| LVESV | 0.395 | 0.182 | 29.5 |

2DE: 2D echocardiography, CMR: cardiovascular magnetic resonance; CoV: coefficient of variance; EDV/ESV: end-diastolic/-systolic volume; EF: ejection fraction; LV: left ventricular; STEMI: ST-segment elevation myocardial infarction; SV: stroke volume.

**Suppl. Table 2.** Correlation analyses between 2D and 3D measurements from CMR 5 days after pPCI for STEMI. Results underlined that LVEF assessment can be performed using either 2D or gold standard 3D CMR. SV assessment should be performed within one dimension due to higher CoV. r indicates the correlation values as obtained from Pearson correlation or r_s_ from Spearman correlation (marked with #) for not normally distributed data.

| **2D CMR vs. 3D CMR** | **r/$r_s$** | **p** | **CoV [%]** |
| --- | --- | --- | --- |
| LVEF# | 0.805 | **<0.001** | 8.8 |
| SV | 0.781 | **<0.001** | 14.6 |
| LVEDV | 0.897 | **<0.001** | 11.3 |
| LVESV# | 0.922 | **<0.001** | 16.6 |

2DE: 2D echocardiography, 3DE: 3D echocardiography; CMR: cardiovascular magnetic resonance; CoV: coefficient of variance; EDV/ESV: end-diastolic/-systolic volume; EF: ejection fraction; LV: left ventricular; STEMI: ST-segment elevation myocardial infarction; SV: stroke volume.

## Supplemental Material

### Expanded Methods

#### Cineventriculography

CVG was performed in 82 patients (*Figure 1*). Standard biplane cineventriculography was obtained using a 30° right anterior oblique (RAO) projection and a 60° left anterior oblique (LAO) projection with injection of at least 20 mL of contrast medium at a flow rate of 14 mL/s using 6F pigtail catheters. In 30 patients, only monoplane CVG in RAO projection was available. As results of the comparison of only monoplane or only biplane CVG to echocardiography or CMR yielded similar results (LVEF similar in all modalities, values of SV, LVEDV and LVESV similar in monoplane or biplane CVG as compared to echocardiography and smaller as compared to CMR) monoplane and biplane measurements were taken as one group.

#### Cardiovascular magnetic resonance

CMR imaging was performed on a 1.5T MR imaging system (Achieva, Philips Healthcare, Best, The Netherlands) using a 32-channel phased array coil within 1 and 5 days after pPCI. Standard cine steady-state-free precession (SSFP) images in standard long-axis geometries (two-, and four-chamber view) on all occasions as well as in short-axis orientation with full ventricle coverage from basis to apex on day 5 after pPCI were acquired for the determination of LV function, volume and shape. Sequence parameters were: repetition time (TR)/echo time (TE) = shortest, flip angle (FA) = 60°, reconstructed voxel size/spatial resolution = 8 × 1.2 × 1.2 mm^3^, phases: 30, one breathhold per slice.

## Notes

**Conflict of interest:** none declared

**Funding:** This work was supported by the Deutsche Forschungsgemeinschaft (DFG, German Research Foundation) - CRC 1116 - (project B05 to C.J., project B11 to A.P., project B12 to M.R. and M.K.)

### Competing Interest Statement

The authors have declared no competing interest.

### Clinical Trial

NCT03539133

### Funding Statement

This work was supported by the Deutsche Forschungsgemeinschaft (DFG, German Research Foundation) - CRC 1116 - (project B05 to C.J., project B11 to A.P., project B12 to M.R. and M.K.)

### Author Declarations

Ethics approval was given by the local ethics committees (Heinrich-Heine-University, Duesseldorf, Germany) complying with the Declaration of Helsinki

