## Supplementary figures and images for "Consistency of left ventricular ejection fraction measurements in the early time course of STEMI"

### Supplemental Figure 1

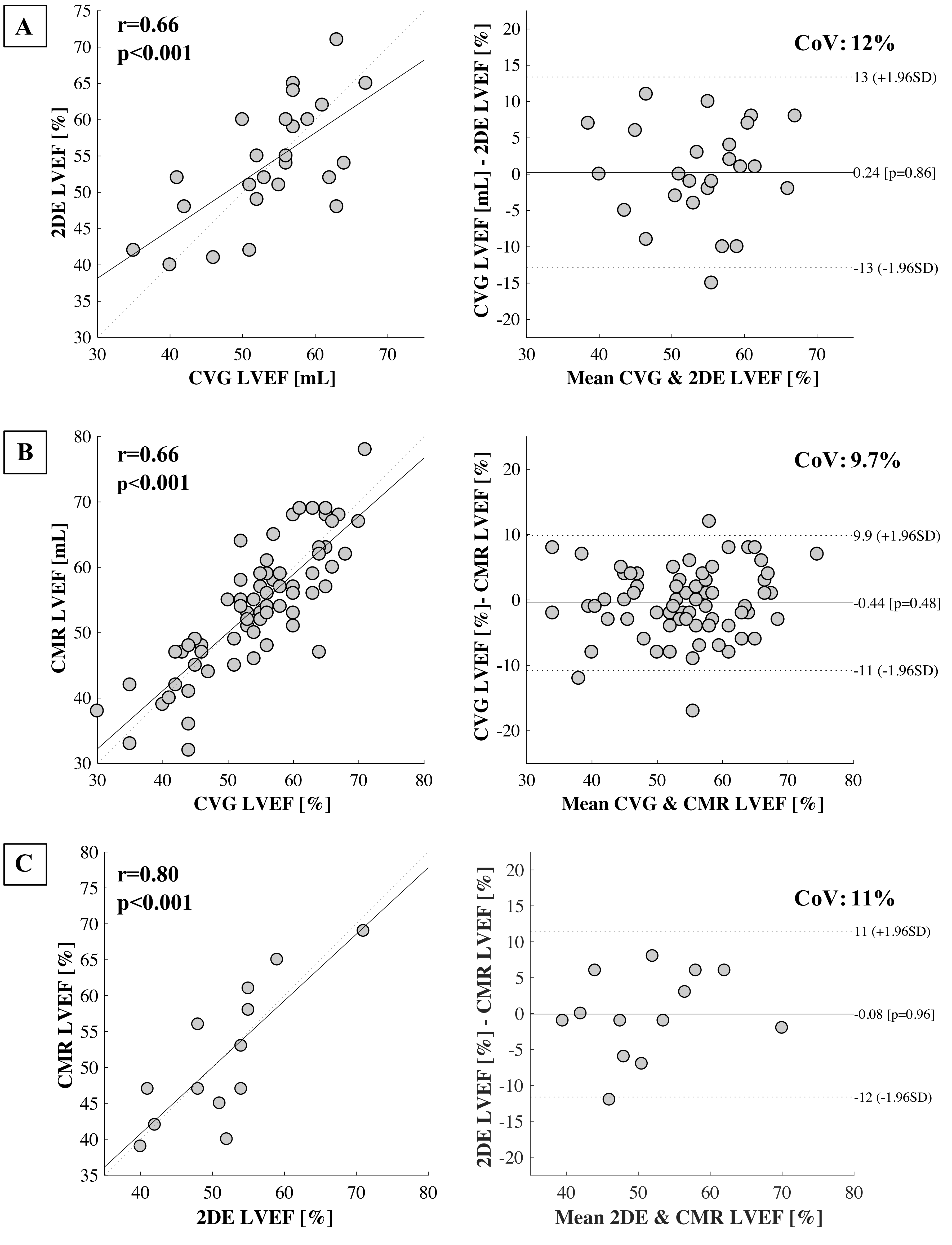

### Supplemental Figure 2

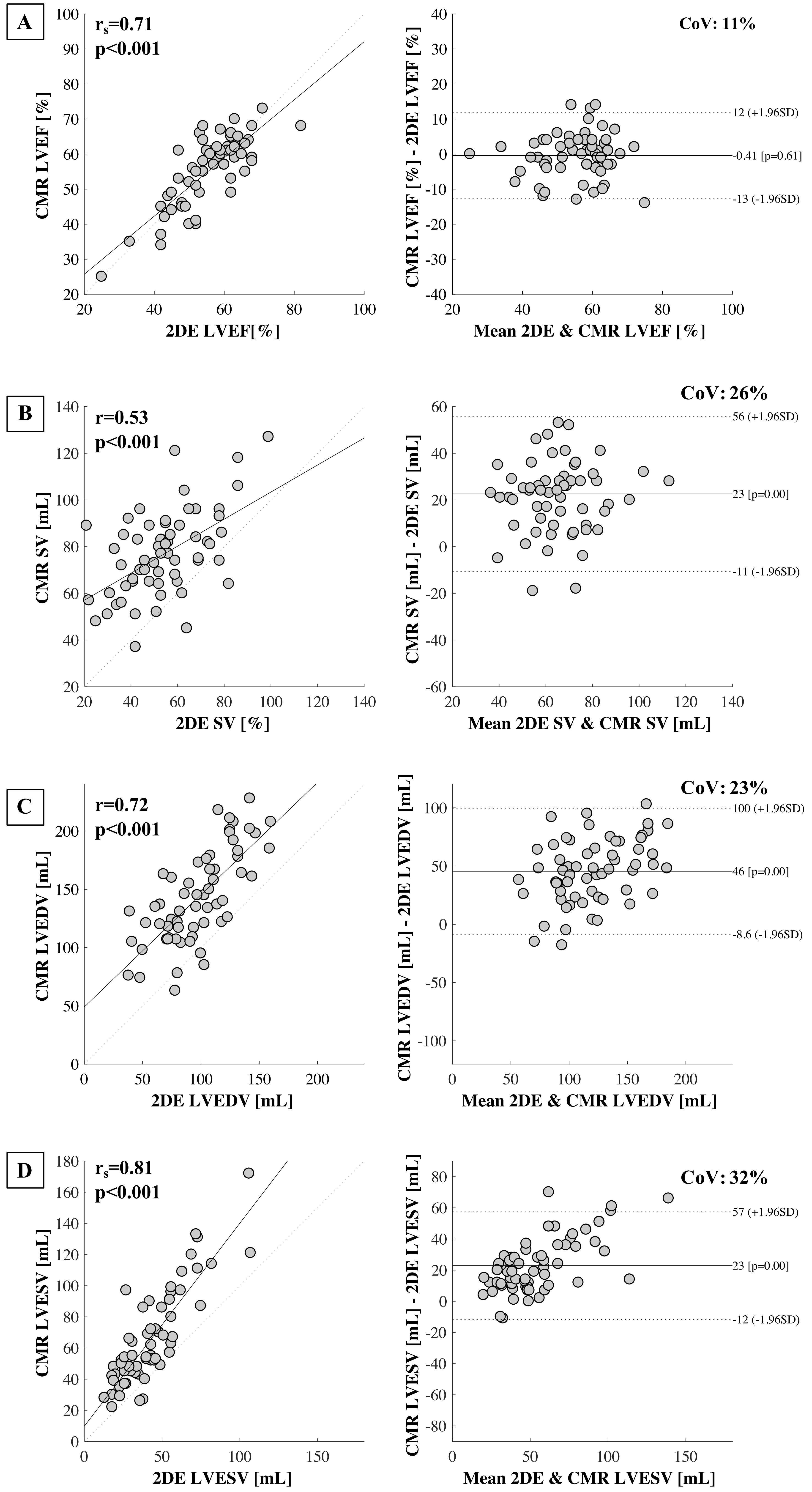
